# Construction of a register-based severity index for anorexia nervosa in Denmark: Association with overall and cause-specific mortality

**DOI:** 10.1101/2025.02.14.25322233

**Authors:** Janne Tidselbak Larsen, Hannah Chatwin, Ziping Zhang, Loa Clausen, Cynthia Marie Bulik, Laura Marie Thornton, Nadia Micali, Liselotte Vogdrup Petersen, Zeynep Yilmaz

## Abstract

**Background:** Considering the high likelihood of chronicity, it is imperative to understand the risk factors and outcomes associated with severe anorexia nervosa (AN), for which Danish registers provide a unique opportunity. We developed a measure of AN severity adapted from clinical literature for use in register-based research.

**Methods:** The study population included all Danish individuals born between 1963 and 2007 who were diagnosed with AN from 1969 to 2013. Using register data, we constructed the Anorexia Nervosa Register-based Severity Index (AN-RSI), incorporating early or late illness onset, number of inpatient admissions and outpatient treatments, cumulative treatment length, and illness duration, each weighted based on clinical importance. Associations between AN-RSI scores, evaluated five years after first AN diagnosis, and mortality were estimated using survival analysis.

**Results:** Among 9,167 individuals diagnosed with AN, 132 died during follow-up: 17 from AN; 30 from suicide; and 85 from other causes. Higher AN-RSI scores were associated with increased rates of mortality from AN, somatic anorexia diagnosis, suicide, alcohol-related causes, and any cause. AN cases who scored in the top 20% of AN-RSI had especially high mortality rates. Furthermore, severe AN cases were also more likely to be in treatment in the next five years after severity was established.

**Conclusions:** AN-RSI effectively captures mortality and long-term treatment in the absence of detailed patient records and is associated with later mortality in AN patients. AN-RSI could serve as a tool to examine epidemiological and genetic risk factors associated with AN course and outcomes.

## Introduction

Anorexia nervosa (AN) is a severe psychiatric disorder characterized by low body weight, severe food restriction, and cognitive distortions related to eating, weight, and shape. Despite some advances in treatment, the prognosis for many individuals with AN remains poor. End-of-treatment remission rates vary between 23-33% for adolescents and 0-25% for adults (Murray *et al*., 2018). Approximately 40% of individuals with AN achieve recovery or remission, but relapse is also common (Fichter *et al*., 2017, Miskovic-Wheatley *et al*., 2023).The impact of AN extends beyond the immediate health consequences, as individuals with AN experience high levels of disability, face unemployment, and frequently receive public assistance (Streatfeild *et al*., 2021). AN is associated with significantly increased mortality (van Eeden *et al*., 2021), with a recent Danish study reported a three-fold increase in overall mortality rates (Larsen *et al*., 2024). The most common causes of death are medical complications related to AN and suicide (Arcelus *et al*., 2011, Larsen *et al*., 2024).

Although severity is a critical issue, there is no consensus definition of severe and enduring AN (SE-AN) (Wonderlich *et al*., 2020). A systematic review found that illness duration and the number of previously failed treatment attempts are the most prevalent criteria across proposed SE-AN definitions (Broomfield *et al*., 2017). Classification guidelines by Hay and Touyz define SE-AN by unrelenting symptoms including dysfunctional behaviors and cognitions associated with AN leading to persistent low body weight, an illness duration of a minimum of three years, and exposure to at least two evidence-based treatments for AN (Hay and Touyz, 2018). Another review found a lack of consistency in the identification of patients with SE-AN, but one of the most commonly used criteria was an illness duration of at least seven years (Ramsay *et al*., 2024). However, the duration criterion is debated due to the lack of consensus on defining chronic AN (Tierney and Fox, 2009) and limited evidence for staging the illness solely by duration (Fernández-Aranda *et al*., 2021, Maguire *et al*., 2008, Maguire *et al*., 2017, Maguire *et al*., 2012). Moreover, greater levels of illness severity and duration may not necessarily be associated with poorer treatment outcomes (Raykos *et al*., 2018). Finally, the term “enduring” is viewed by some as precluding the possibility of recovery and recommend the term “longstanding” instead.

Denmark is uniquely positioned to study the epidemiology of AN thanks to its comprehensive population registers. The use of population-based samples minimizes selection bias and accurately represents cases as detected throughout the national healthcare system. This allows for robust longitudinal studies, which are essential for understanding the progression and outcomes of AN over time (Chatwin *et al*., 2023, Larsen *et al*., 2024, Momen *et al*., 2022). Despite the advantages of these registers, detailed clinical records are currently not readily available and only have become electronic in recent years. Given the high morbidity and mortality associated with AN and the lack of a consensus definition of SE-AN, a practical and standardized method to capture AN severity using register-based data could prove valuable. In addition to a binary classification system, access to rich health data from Danish registers provides a unique opportunity to develop a more comprehensive and nuanced *continuous* measure of severity while incorporating some of the key aspects of the classification guidelines.

The aim of this study was to construct a register-based severity index for AN, which could facilitate the identification of severe AN cases in Denmark and countries with similar data sources and contribute to a better understanding of the etiology and course of AN. The secondary aim of this study was to evaluate the associations between the severity index and overall and cause-specific mortality in individuals with AN.

## Methods

### Sources

This study uses data from multiple nationwide Danish registers. The Danish Civil Registration System (Pedersen, 2011) contains information on date of birth and vital status of all individuals who have been alive and had permanent residence in Denmark since 1968. The Danish National Patient Register (Lynge *et al*., 2011) have recorded inpatient admission to somatic hospital wards since 1977, and the Danish Psychiatric Central Research Register (PCRR) (Mors *et al*., 2011) have recorded inpatient admission to psychiatric wards since 1969. Since 1995, NPR and PCRR have also recorded outpatient contacts to hospital. The Danish Register of Causes of Death (Helweg-Larsen, 2011) has recorded date and main and contributing causes of death since 1970.

The International Classification of Diseases, 8^th^ Revision (ICD-8) was used in Denmark until 1994, when it was replaced by the International Classification of Diseases, 10^th^ Revision (ICD-10).

### Anorexia Nervosa Register-based Severity Index

AN severity was measured using an Anorexia Nervosa Register-based Severity Index (AN-RSI), calculated as the sum of points assigned to each of the included severity-related variables available in the Danish registers. These six variables (described below) were identified by previous studies, and the points and weights were assigned with input from experienced clinicians. For the main cohort, the AN-RSI sum score was calculated five years after date of first diagnosis. Given the lack of consensus in the literature concerning duration, with studies often using follow-up periods of three or seven years (Hay and Touyz, 2018, Ramsay *et al*., 2024), we chose the intermediate period of five years to balance the need for sufficient data to capture illness progression and practical considerations of duration of follow-up and potential attrition. Overall, the aim was to provide a measure for AN severity that is robust, practical, and clinically relevant.

The six severity-related variables included in the AN-RSI were as follows:

*1. Early onset* was defined as age below 10 years at start of first in- or outpatient hospital contact with AN diagnosis. Assigned 1 point.
*2. Late onset* was defined as age above 25 years at start of first in- or outpatient hospital contact with AN diagnosis. Assigned 1 point.
*3. Inpatient admissions* were defined as the number of recorded inpatient hospital admissions with AN diagnosis. Assigned 2 points per admission.
*4. Outpatient treatments* were defined as the number of recorded outpatient treatment courses with AN diagnosis, disregarding the first treatment course. Assigned 1 point per treatment.
*5. Treatment length* was defined as the combined lengths of inpatient and outpatient treatment. The length of an inpatient admission was defined as the difference between start and end dates. The length of an outpatient treatment was defined as the sum of the difference between start and end dates and the recorded number of treatment days (days with an appointment) during the outpatient treatment course, both multiplied by 0.5.
*6. Illness duration* was defined as time since date of first AN diagnosis until the latest treatment day occurring during the five years after first diagnosis for the main definition or until each treatment time point (i.e., start and end dates of each hospital contact) for the continuously updated definition used in sensitivity analysis. Assigned 1 point per full year.

### Sensitivity analyses

For the expanded cohort with full follow-up, AN-RSI was continuously updated at each treatment time point until 31 December 2018.

For additional sensitivity analyses, we: (1) examined the individual AN severity variables; (2) performed sex-stratified analysis; and (3) defined alternative versions of the individual AN severity variables and the combined AN-RSI using only inpatient admission data in all definitions of AN cases and measures of AN severity (due to the outpatient hospital contact information not being available before 1995).

### Study population

The main cohort comprised all individuals living in Denmark, born after 1 January 1963 and 31 December 2007, and diagnosed with AN (ICD-8: 306.50; ICD-10: F50.0, F50.1) during an in- or outpatient contact to hospital after their sixth birthday and between 1 January 1969 and 31 December 2013. The F50.1 (atypical AN) diagnostic code was included to account for individuals with symptoms similar to AN but miss one symptom from F50.0 diagnosis.

These are often mild cases where the person has not lost sufficient weight, or more severe cases where the person is significantly underweight but does not endorse the psychological symptoms. These criteria and timeline enabled us to evaluate AN severity five years after the first diagnosis for all included individuals and until 31 December 2018 at the latest. Those who died or emigrated during the first five years after onset were excluded.

### Sensitivity analysis

To gain power and include as much as the available data as possible, the cohort was expanded to include those diagnosed after their sixth birthday, not limited to specific calendar years. For this cohort, AN severity was evaluated and updated continuously at each new hospital contact during follow-up from the date of the first diagnosis until death, emigration, or 31 December 2018, whichever came first.

### Outcomes

The main outcome was mortality from any cause. Secondary outcomes were mortality from eight specified causes of death: (1) AN [ICD-8: 306.5; ICD-10: F50.0, F50.1]; (2) other eating disorders [ICD-10: F50.2, F50.3, D50.8, F50.9]; (3) somatic anorexia diagnosis [ICD-10: R63.0]; (4) suicide [ICD-8: 950-959; ICD-10: Y87.0, X60-X84]; (5) accident or intention unknown [ICD-10: T96, X40-X49, Y10-Y19, Y40-Y59, F1x.0]; (6) alcohol-related causes [ICD-10: K70, F10.2 (unless main cause of death is listed as X09)]; (7) other specified psychiatric diagnoses [ICD-10: F00-F09, F20-F99, F10-F19 (excluding F1x.0)]; and (8) other causes of death [any not listed above]. This list was compiled and causes of death were classified accordingly after a thorough review of all causes of death for everyone with an AN diagnosis by the senior author (ZY) and approved by two co-authors who are clinicians (LC and CB). We included somatic anorexia diagnosis as a separate cause of death based on the diagnostic terminology in Danish (*anoreksi*), as this difference may have resulted some healthcare providers to select this code over the diagnostic code for AN. These eight causes of death were defined in a hierarchical manner in the order listed above, so that each individual was included in only one cause-of-death group.

### Statistical analysis

We conducted survival analyses using Cox regression resulting in hazard ratios (HR) and 95% confidence intervals (CI) for associations between AN-RSI and mortality, adjusted for time since first AN diagnosis as the continuous underlying time scale, and calendar year of first diagnosis and birth year in 10-year strata.

For the main analysis, follow-up started five years after the first diagnosis, at which point AN-RSI was also evaluated. Follow-up ended at death, emigration, or on December 31, 2018, whichever came first.

The results of AN-RSI were evaluated in two ways: (1) the raw continuous index score resulting in HR per one point increase in AN-RSI; and (2) severe AN cases—defined as individuals who scored in the top 20% of AN-RSI scores five years after initial diagnosis — compared to less severe AN cases in the bottom 80% of AN-RSI. This cut-off is in line with previous studies showing that approximately 20% of individuals with AN do not improve with treatment (Dalle Grave, 2020).

We calculated the proportion of severe versus less severe AN cases receiving any in- or outpatient treatment for AN annually for up to 10 years post-diagnosis. Additionally, we assessed the prevalences of diagnosed psychiatric comorbidities prior to first AN diagnosis, during the severity evaluation period (0-5 years post-diagnosis), and in the subsequent five years (5-10 years post-diagnosis) for severe and less severe cases. Details on the definitions and groupings of the included psychiatric conditions can be found elsewhere (Momen *et al*., 2022). Statistically significant differences between severe and less severe AN cases were evaluated using Pearson’s chi-squared tests.

### Sensitivity analyses

For the analysis of the expanded cohort, follow-up started at first AN diagnosis, and AN-RSI was continuously updated throughout follow-up and treated as a time-dependent exposure. Follow-up ended at death, emigration, or on December 31, 2018, whichever came first. The same severity threshold value from the main analysis was used to define severe AN cases.

For the main cohort, we analyzed the association between each severity variable included in AN-RSI and overall mortality. Results are presented per one point increase and for severe compared to less severe AN cases. We also conducted sex-stratified analyses of the association between AN-RSI and overall mortality in the main cohort. The small number of men with diagnosed AN precluded further examination of the associations between AN-RSI and specific causes of death.

Finally, we conducted analyses on the alternative definition of AN-RSI using only inpatient admission data in the main cohort.

## Results

A total of 9,167 individuals [8,592 females (93.73%), 575 males (6.27%)] were included in the main analysis. The distribution of severity variables contributing to the combined AN-RSI is shown in **Supplementary Figure 1**. The main contributors were number of inpatient admissions, number of outpatient treatments, and illness duration.

Based on AN-RSI, 2,036 individuals (22.21%) were classified as severe AN cases (scoring 6 points or more) five years after onset, of whom 96 (4.72%) were male. **Figure 1** shows the proportion of those still followed in the survival analysis who are in treatment annually up to 10 years from first AN diagnosis (not counting the date of onset in the first year) for severe and less severe AN cases, respectively. We found that a larger proportion of AN cases categorized as severe five years after first diagnosis were more likely to be in treatment during the first five years, as would be expected based on the construction of the AN-RSI. Importantly, severe AN cases were also more likely to be in treatment in the next five years after they had attained the threshold for being considered severe. **Table 1** shows the number of diagnosed psychiatric conditions during each time period for severe and less severe AN cases, while **Supplementary Table 1** shows the proportions diagnosed with each psychiatric condition. Severe AN cases also had significantly higher prevalences of comorbid psychiatric conditions during and after the severity evaluation period compared to less severe cases, with the exception of anxiety disorders and other eating disorders having a higher prevalence in severe AN cases also prior to first AN diagnosis.

**Figure 1.**
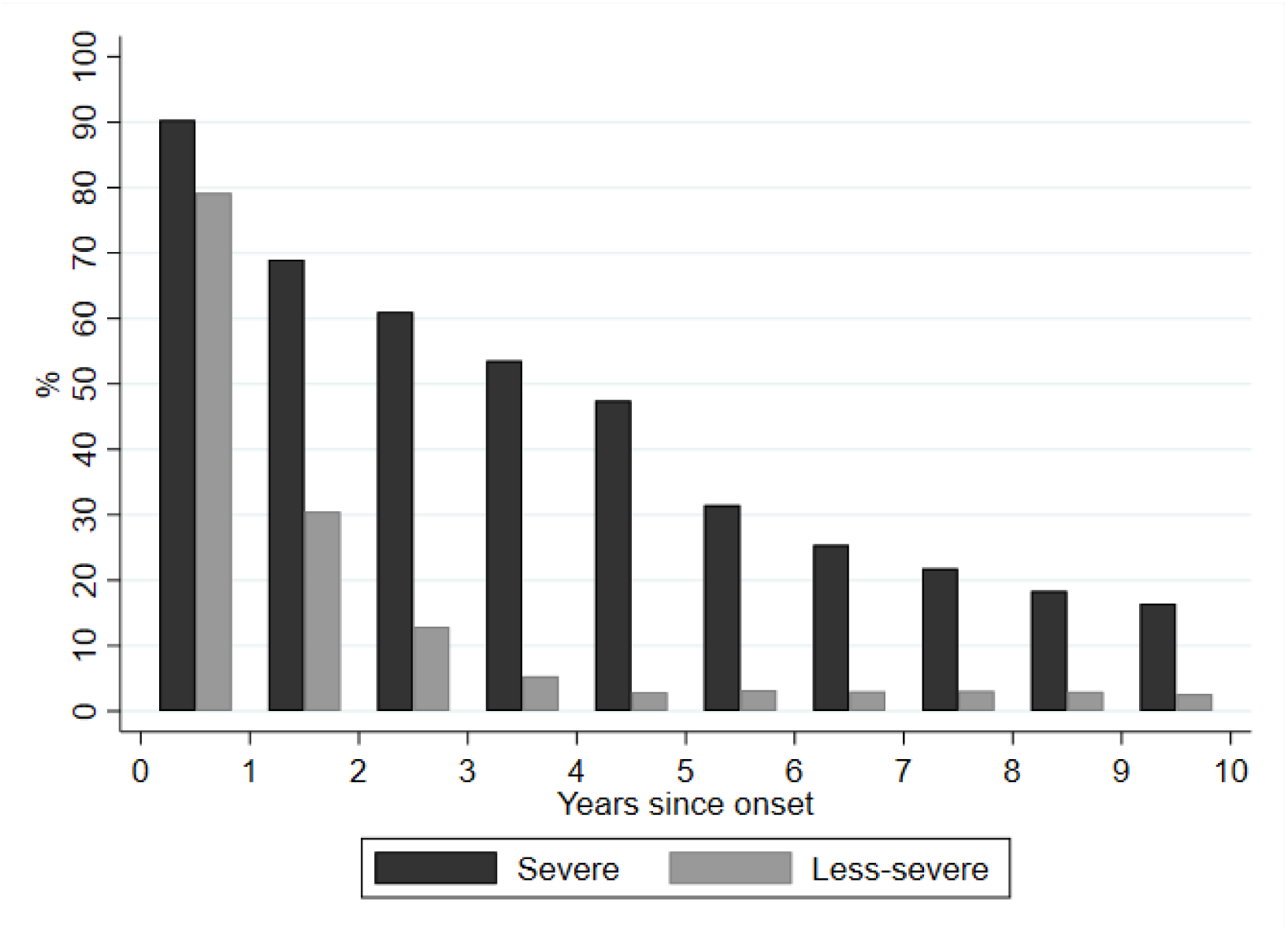
Severe and less-severe anorexia nervosa cases (evaluated 5 years after onset) in treatment during each year since onset, calculated as the proportion of those still followed in the survival analysis.

**Table 1.** Number of comorbid psychiatric conditions diagnosed prior to AN diagnostic onset; during severity evaluation (years 0-5 after AN onset); and during the first 5 years after severity evaluation (i.e. years 5-10 after AN onset) for severe and less severe AN cases.

|  |  | Less severe AN<br>N=7,131 |  | Severe AN<br>N=2,036 |  | p |
| --- | --- | --- | --- | --- | --- | --- |
|  | Number of comorbidities | n | % | n | % |  |
| Prior to AN onset | 0 | 5,204 | 72.98 | 1,436 | 70.53 | 0.060 |
|  | 1 | 1,013 | 14.21 | 295 | 14.49 |  |
|  | 2 | 498 | 6.98 | 153 | 7.51 |  |
|  | 3 | 257 | 3.60 | 93 | 4.57 |  |
|  | 4 | 116 | 1.63 | 38 | 1.87 |  |
|  | 5+ | 43 | 0.60 | 21 | 1.03 |  |
| During severity evaluation | 0 | 2,497 | 35.02 | 428 | 21.02 | <0.001 |
|  | 1 | 2,421 | 33.95 | 595 | 29.22 |  |
|  | 2 | 1,238 | 17.36 | 448 | 22.00 |  |
|  | 3 | 574 | 8.05 | 296 | 14.54 |  |
|  | 4 | 270 | 3.79 | 169 | 8.30 |  |
|  | 5+ | 131 | 1.84 | 100 | 4.91 |  |
| After severity evaluation | 0 | 5,516 | 77.35 | 1,115 | 54.76 | <0.001 |
|  | 1 | 749 | 10.50 | 386 | 18.96 |  |
|  | 2 | 488 | 6.84 | 290 | 14.24 |  |
|  | 3 | 264 | 3.70 | 144 | 7.07 |  |
|  | 4 | 86 | 1.21 | 64 | 3.14 |  |
|  | 5+ | 28 | 0.39 | 37 | 1.82 |  |

We observed 132 deaths during the follow-up period. Of the AN cases classified as severe, 46 cases (2.26%) died, while 86 (1.21%) of the less severe AN cases died. **Figures 2-3** show the associations between AN-RSI and overall and cause-specific mortality per one point increase in AN-RSI score and for severe compared to less severe AN cases, respectively. In addition to mortality from any cause [HR=1.03 (95% CI: 1.02, 1.04)], causes of mortality most strongly associated with increase in AN-RSI score were AN [1.07 (1.04, 1.10)], somatic anorexia [1.07 (1.02, 1.11)], other psychiatric disorders [1.07 (1.03, 1.11)], and suicide [1.02 (1.00, 1.04)]. The only outcome negatively associated with increase in AN-RSI score was mortality from other eating disorders, although these results were based on few deaths (n=6) and not statistically significant [0.95 (0.79, 1.14)]. In the categorical analysis, the HR of mortality from any cause was 1.92 (1.34, 2.75) for severe AN cases compared to less severe cases, and being classified as a severe case was significantly associated with mortality from AN [6.74 (2.49, 18.24)], anorexia (somatic diagnosis) [4.62 (1.23, 17.33)], and alcohol-related causes [3.58 (1.03, 12.44)].

**Figure 2.**
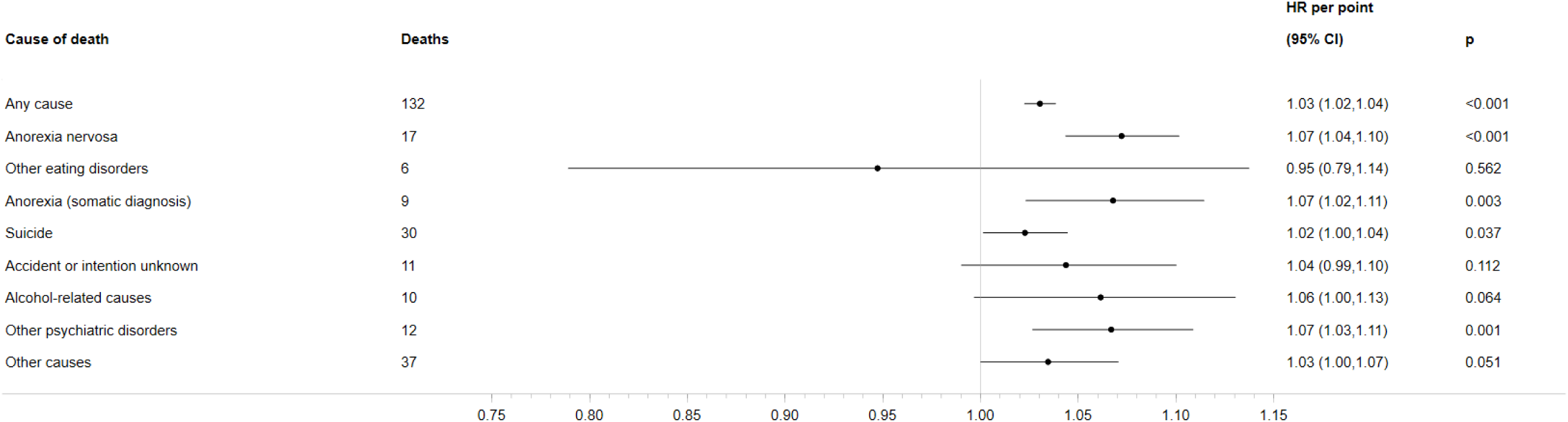
Hazard ratios (HR) with 95% confidence intervals (CI) for overall and cause-specific mortality per one point increase of the Anorexia Nervosa Register-based Severity Index evaluated 5 years after onset.

**Figure 3.**
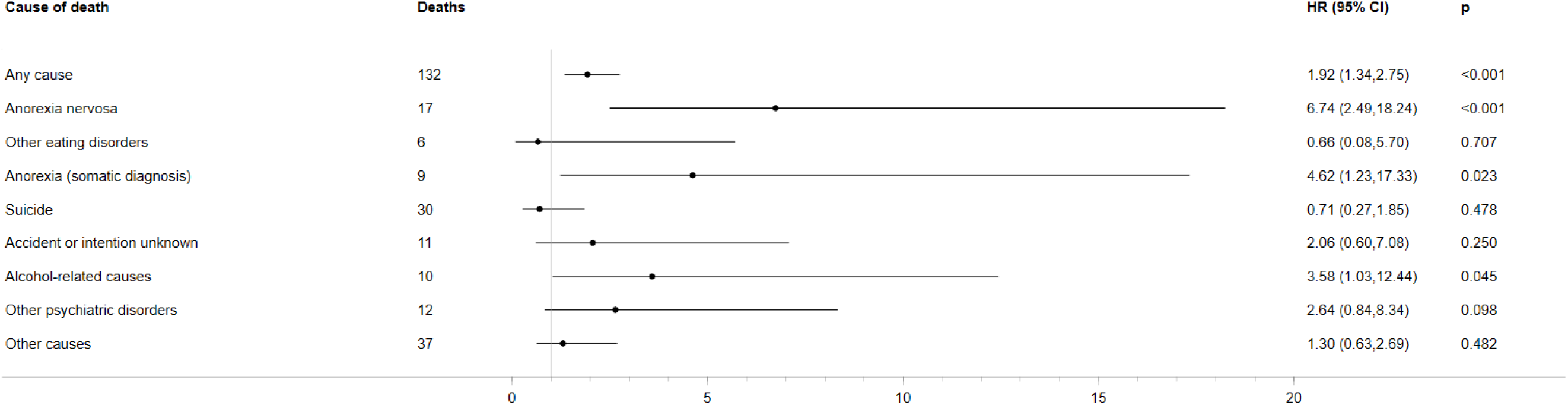
Hazard ratios (HR) with 95% confidence intervals (CI) for overall and cause-specific mortality for severe anorexia nervosa cases compared to less-severe anorexia nervosa cases evaluated 5 years after onset.

### Sensitivity analyses

**Supplementary Figure 2** shows the distribution of severity measures contributing to AN-RSI at the end of follow-up for the expanded cohort with continuously updated AN-RSI scores.

Compared to the five-year severity evaluation, illness duration contributed more to the combined score since no upper time limit of illness duration was imposed. With the expanded cohort of 13,833 [12,932 females (93.49%), 901 males (6.51%)] and longer follow-up, we observed 243 deaths during follow-up. 22.80% of individuals included in the cohort and 39.51% of those who died were categorized as severe cases. HRs of mortality from any cause was 1.03 (95% CI: 1.02, 1.03) per one point increase. Mortality from all included causes of death except suicide were significantly associated with per-point increase in AN-RSI (**Supplementary Figure 3**). Comparing severe to less severe AN cases, the HR of mortality from any cause was 3.07 (2.32, 4.05), and being classified as a severe AN case was significantly associated with mortality from AN, somatic anorexia, alcohol-related causes, and accident or intention unknown (**Supplementary Figure 4**).

In the main cohort, the individual severity variables measured at five years capturing late onset, number of inpatient admissions, number of outpatient treatments, and illness duration, were all significantly associated with mortality from any cause, whereas treatment length was not. The distribution of points generated by the early onset criterion did not allow for estimation (**Supplementary Figure 5**). Severe AN classification solely based on the number of inpatient admissions or illness duration was significantly associated with mortality from any cause (**Supplementary Figure 6**).

Of the 132 deaths from any cause in the main analysis, eight were male. Only the results based on females generated significant associations between mortality from any cause and AN-RSI and comparing severe to less severe AN cases. However, the point estimates from male-only analyses were higher in all three instances (**Supplementary Figure 7**).

Using only inpatient admission data to define AN cases and AN-RSI meant that the vast majority of AN-RSI evaluated at five years was derived from number of inpatient admissions (**Supplementary Figure 8**), and the cohort was limited to 3,464 AN cases [3,223 females (93.04%), 241 males (6.96%)]. We observed 86 deaths during follow-up. Due to low counts in some cause-of-mortality groups, we were unable to estimate associations for these causes of mortality or report number of deaths due to each specific cause to comply with Danish legislature on person-identifiable register information. In general, the results were similar to the main results but with wider CI due to the smaller cohort (**Supplementary Figures 9-10**).

## Discussion

This study is the first to construct a register-based severity index as a proxy for AN severity in the absence of detailed clinical records. The development of AN-RSI addresses a significant gap in the field, especially given the limited treatment options and inconsistency in treatment approaches for the potentially fatal SE-AN (Wonderlich *et al*., 2024, Wonderlich *et al*., 2020). Our findings indicate that both higher AN-RSI scores and the classification of severe AN cases were associated with overall mortality. However, the specific causes of mortality significantly associated with severity varied depending on whether the continuous AN-RSI or the categorical classification of AN cases was used. The continuous AN-RSI was associated with increased mortality from AN, somatic anorexia, other psychiatric disorders, and suicide. In contrast, classification of severe AN cases was not significantly associated with suicide or other psychiatric disorders. Instead, severe cases had higher rates of death from AN, somatic anorexia, and alcohol-related causes. These distinctions highlight the importance of considering both a continuous measure of AN severity and a binary severity classification when assessing prognoses and outcomes. That these specific causes of mortality are associated with AN-RSI suggests that AN severity is closely linked to underlying mental health problems and behaviors directly related to the disorder, and that severe AN may exacerbate vulnerabilities to other psychiatric disorders, alcohol use, and suicidal ideation, which in turn increase mortality risk. These findings highlight the need for effective treatment approaches that address not only AN itself but also a broader spectrum of mental health problems and behaviors associated with severe AN. Notably, death from other eating disorders was not associated with AN severity, suggesting that the factors driving mortality in severe AN are distinct and do not apply to individuals with AN who diagnostically transition to another eating disorder.

The analysis of the expanded cohort and including the AN-RSI continuously updated throughout the follow-up period yielded similar results to the main analyses using AN-RSI evaluated at five years after AN onset, indicating the robustness of our findings. The expanded cohort and extended evaluation period provided greater statistical power, but the five-year post-onset AN-RSI offers a standardized time frame that facilitates comparison across individuals and is informed by the existing literature.

Each individual severity variable was associated with overall mortality except treatment length (analysis not available for early onset), providing evidence for treatment contact reducing the risk of mortality in longstanding cases of AN. This finding also emphasizes the need for early intervention for patients with AN and close monitoring of those with frequent hospital contacts and longer illness duration to reduce mortality risk. It also suggests that individuals who are diagnosed with AN later in life may face higher risks, possible due to delayed diagnosis and comorbidity (Søeby *et al*., 2025).

The results pertaining to mortality were mainly driven by females, as very few men (n=8) with an AN diagnosis died during follow-up. Consequently, we did not obtain significant results of overall mortality from male-only analyses. This sex disparity highlights the need for further research to understand the impact of AN severity in males.

To address the limitation of outpatient contact data being unavailable until 1995, we constructed the alternative AN-RSI using only inpatient contacts, ensuring that AN-RSI was uniformly defined throughout the observation period. The results from this alternative AN-RSI were similar to those derived from AN-RSI using both in- and outpatient contacts. This consistency suggests that the original AN-RSI is robust and reliable, even in the absence of outpatient information for parts of the observation period, and it remains a valid tool for assessing AN severity across different time frames.

AN cases classified as severe based on AN-RSI five years after first diagnosis continued to exhibit higher rates of any type of treatment for AN in the five years after they had attained the threshold for being considered severe. Although the construction of the AN-RSI did not specifically focus on long-standing AN, as ongoing AN diagnoses in the registers during or after the five-year severity establishment period were not required, the sustained need for treatment underscores the chronic nature of severe AN. Combined with the increased prevalences of psychiatric comorbidities before and after onset of AN, this finding shows that AN-RSI captures several important aspects of illness course.

Of note, AN-RSI captured a higher psychiatric comorbidity burden in individuals with severe versus less severe AN. This further reflects the robustness of the AN-RSI as comorbidity has often been associated with severity despite not being part of the primary definition (Hay and Touyz, 2015, Wildes *et al*., 2017). More specifically, we observed higher prevalence of comorbid psychiatric conditions after AN diagnosis, before and after the period in which we assessed severity. One possible explanation is that contact with the healthcare system for AN diagnosis may have resulted in assessment and diagnosis of other psychiatric disorders (Berkson, 1946). Overall, all psychiatric diagnoses—grouped based on ICD chapters—were more commonly diagnosed in severe cases compared to less severe cases. However, only anxiety and other eating disorders were more prevalent in severe AN cases prior to first AN diagnosis, which reflects the large body of research on AN risk factors (Abdulkadir *et al*., 2025, Bulik *et al*., 1997, Halmi *et al*., 2005, Kerr-Gaffney *et al*., 2018, Keski-Rahkonen and Mustelin, 2016, Schaumberg *et al*., 2019, Wildes *et al*., 2017). Taken together, our findings highlight the important relationship between AN-based severity and the higher burden of comorbid psychiatric disorders.

### Strengths and limitations

Our study has several strengths that enhance the robustness and reliability of our findings. Firstly, the use of a large cohort provides substantial statistical power and generalizability to our results. The comprehensive registers offer rich and detailed information, allowing for thorough analysis and accurate classification of AN severity. Additionally, the reliance on register data minimizes recall bias. Furthermore, validity of register-based diagnoses in Denmark has been established for a multitude of psychiatric disorders (Bock *et al*., 2009, Nissen *et al*., 2017, Uggerby *et al*., 2013), including eating disorders (Egedal *et al*., 2024). Eating disorder diagnoses have also been validated in the similar Swedish health registers (Birgegård *et al*., 2022). Furthermore, the public healthcare system in Denmark reduces selection bias and enhance the applicability of our findings to similar healthcare settings.

Our study also has several limitations that should be acknowledged. Firstly, we relied solely on hospital diagnoses and treatments, which may not capture the full spectrum of AN severity, although severe AN cases are much more likely to be captured by the hospital-based diagnoses. The initial hospital diagnosis may also not accurately capture the actual time of onset. Secondly, the weighting of severity variables contributing to the combined AN-RSI was based on clinical judgement, which introduces a degree of subjectivity despite their grounding in the clinical research literature. Additionally, there may be differences in clinicians’ practices regarding use of primary diagnosis for treatment and cause of death, potentially affecting the consistency of our data. For instance, AN cases with personality disorders may have the latter listed as the primary diagnosis for treatment targeting AN since it may be viewed as pervasive; similarly, any somatic reason for treatment directly caused by AN may be entered as the primary diagnosis by a clinician. It is also possible that some deaths from suicide are recorded as accidents, cases of unknown intent, or attributed to other causes due to lack of evidence of intent or a desire to avoid the stigma associated with suicide. As we summarized above, we believe the instances of somatic anorexia as a cause of death may have been due to differences in language-specific diagnostic terminology, and this anomaly would explain the significant associations we observed with somatic anorexia and AN severity as well as severe AN. Moreover, we originally intended to include a cause-of-death category of somatic diagnoses potentially attributable to AN, but the number was too low to warrant its own category. The construction of the AN-RSI was based exclusively on AN-related measures, without incorporating other clinically relevant data such as body mass index, symptom-level data, or questionnaires, which are not available in the registers. The lack of comprehensive clinical data may affect the precision of our severity measures and the generalizability of our results to other settings. Finally, since AN-RSI relies heavily on treatment-seeking variables (i.e., inpatient admissions, outpatient readmissions, and treatment length), we were unable to capture individuals (especially severe AN cases) who are not in contact with the healthcare system or choose not to seek treatment. However, illness duration was included as a variable to account for non-treatment-seeking individuals as best as we can using register data, which were also at least partially captured by non-psychiatric healthcare contacts that may be associated with AN-related complications or medical stabilization.

### Conclusion

Our study demonstrates that AN-RSI is associated with comorbidity and mortality and therefore shows promise in its use and applications in register-based research. AN-RSI provides a method to quantify and measure AN severity in the absence of detailed clinical data, which is crucial given the limited knowledge about the biological and epidemiological risk factors associated with severe AN. Future studies should examine other correlates such as somatic comorbidities, validate AN-RSI in other datasets, add genetic indices such as polygenic scores, and explore how well it correlates with other severity indicators such as body mass index and eating disorder symptoms. It is our expectation that AN-RSI can serve as a tool to examine epidemiological and genetic risk factors associated with the course and outcomes of AN.

## Data Availability

Access to data requires application to the Danish Health Data Authority and the Danish Data Protection Agency.

