## Supplementary Tables and Figures for "Construction of a register-based severity index for anorexia nervosa in Denmark: Association with overall and cause-specific mortality"

**Supplementary Figure 1.** Distribution of severity variables contributing to the combined Anorexia Nervosa Register-based Severity Index evaluated 5 years after onset.

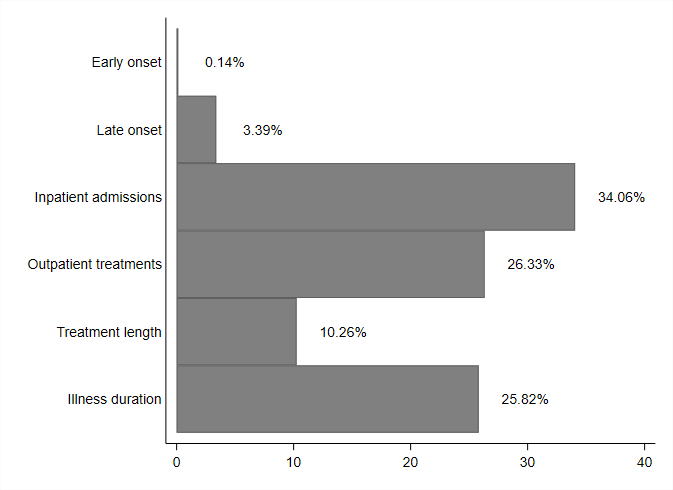

**Supplementary Table 1.** Psychiatric conditions diagnosed prior to AN onset, during severity evaluation (years 0-5 after AN onset), and the first 5 years after severity evaluation (i.e. years 5-10 after AN onset) for severe and less severe AN cases.

|  |  | Less severe AN  N=7,131 | | Severe AN  N=2,036 | | p |
| --- | --- | --- | --- | --- | --- | --- |
|  | **Comorbidity** | **n** | **%** | **n** | **%** |  |
| Prior to onset | Organic | 19 | 0.27 | 6 | 0.29 | 0.829 |
|  | Substance use | 247 | 3.46 | 70 | 3.44 | 0.955 |
|  | Schizophrenia | 146 | 2.05 | 47 | 2.31 | 0.469 |
|  | Mood | 520 | 7.29 | 161 | 7.91 | 0.350 |
|  | Anxiety | 759 | 10.64 | 249 | 12.23 | 0.044 |
|  | Other eating disorders | 975 | 13.67 | 335 | 16.45 | 0.002 |
|  | Personality | 414 | 5.81 | 138 | 6.78 | 0.104 |
|  | Intellectual | 33 | 0.46 | 12 | 0.59 | 0.471 |
|  | Developmental | 71 | 1.00 | 27 | 1.33 | 0.201 |
|  | Behavioral | 291 | 4.08 | 99 | 4.86 | 0.123 |
| During severity evaluation | Organic | 61 | 0.86 | 22 | 1.08 | 0.344 |
|  | Substance use | 441 | 6.18 | 149 | 7.32 | 0.066 |
|  | Schizophrenia | 365 | 5.12 | 230 | 11.30 | <0.001 |
|  | Mood | 1,381 | 19.37 | 626 | 30.75 | <0.001 |
|  | Anxiety | 1,574 | 22.07 | 650 | 31.93 | <0.001 |
|  | Other eating disorders | 2,753 | 38.61 | 1,048 | 51.47 | <0.001 |
|  | Personality | 1,072 | 15.03 | 474 | 23.28 | <0.001 |
|  | Intellectual | 79 | 1.11 | 37 | 1.82 | 0.012 |
|  | Developmental | 190 | 2.66 | 117 | 5.75 | <0.001 |
|  | Behavioral | 480 | 6.73 | 239 | 11.74 | <0.001 |
| After severity evaluation | Organic | 19 | 0.27 | 20 | 0.98 | <0.001 |
|  | Substance use | 250 | 3.51 | 121 | 5.94 | <0.001 |
|  | Schizophrenia | 258 | 3.62 | 175 | 8.60 | <0.001 |
|  | Mood | 528 | 7.40 | 293 | 14.39 | <0.001 |
|  | Anxiety | 683 | 9.58 | 351 | 17.24 | <0.001 |
|  | Other eating disorders | 526 | 7.38 | 435 | 21.37 | <0.001 |
|  | Personality | 516 | 7.24 | 318 | 15.62 | <0.001 |
|  | Intellectual | 30 | 0.42 | 17 | 0.83 | 0.021 |
|  | Developmental | 69 | 0.97 | 46 | 2.26 | <0.001 |
|  | Behavioral | 127 | 1.78 | 74 | 3.63 | <0.001 |

**Supplementary Figure 2.** The distribution of severity variables contributing to the combined Anorexia Nervosa Register-based Severity Index evaluated at end of the full, available follow-up period.

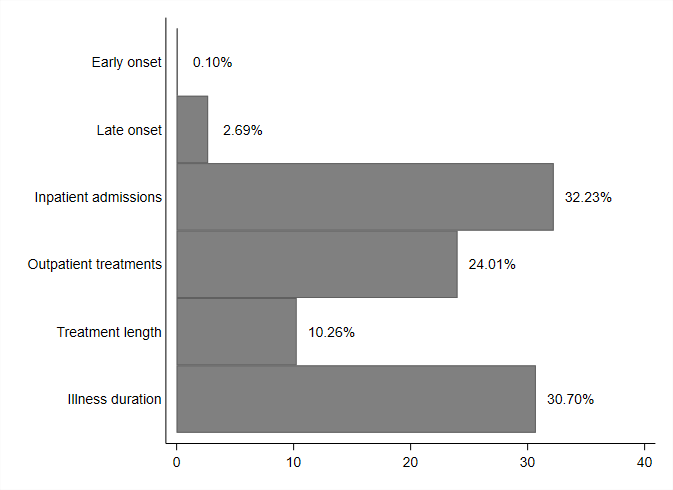

**Supplementary Figure 3.** Hazard ratios (HR) with 95% confidence intervals (CI) for overall and cause-specific mortality per one point increase of the Anorexia Nervosa Register-based Severity Index evaluated continuously throughout the full, available follow-up period.

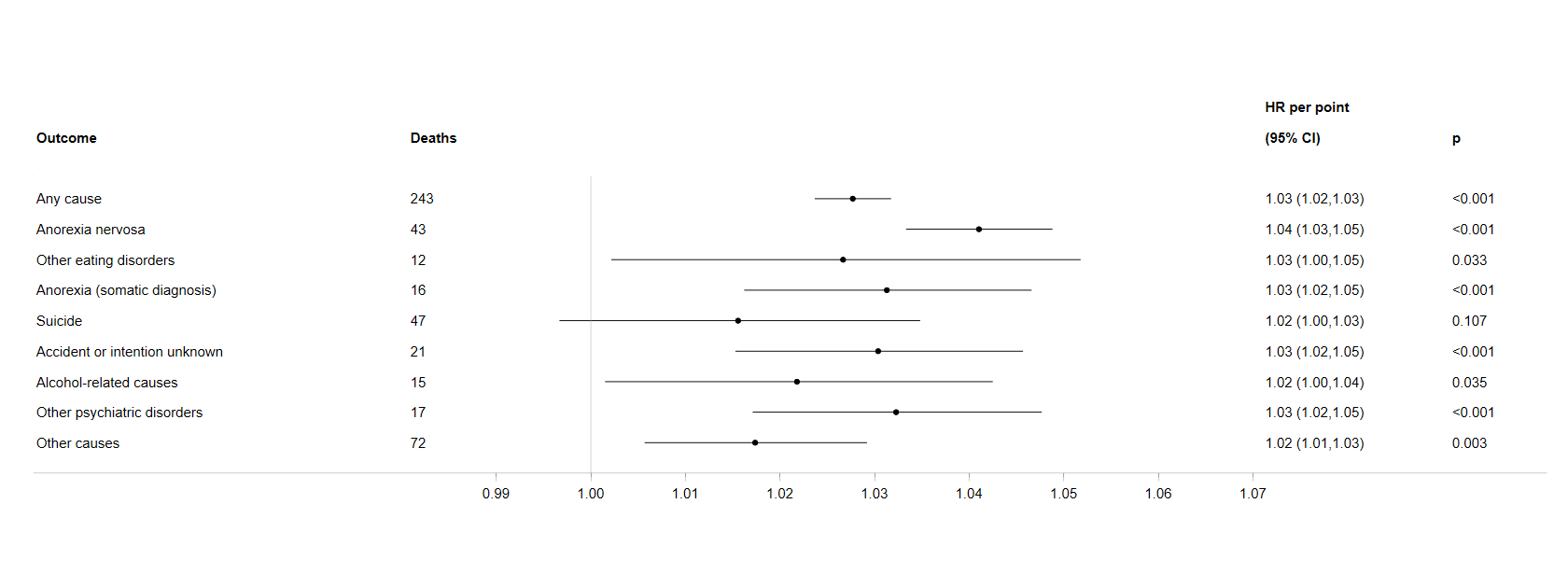

**Supplementary Figure 4.** Hazard ratios (HR) with 95% confidence intervals (CI) for overall and cause-specific mortality for severe anorexia nervosa cases compared to less-severe anorexia nervosa cases evaluated continuously throughout the full, available follow-up period.

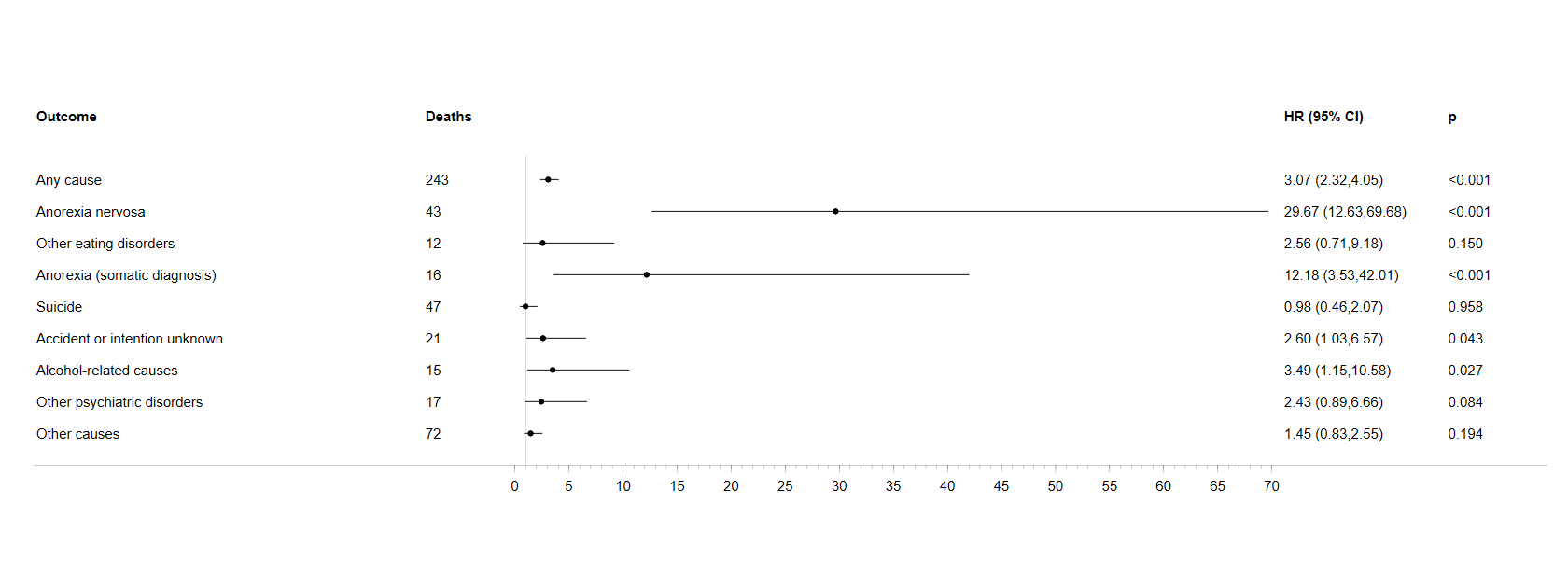

**Supplementary Figure 5.** Hazard ratios (HR) with 95% confidence intervals (CI) for overall mortality per one point increase of each of the individual severity variables contributing to the Anorexia Nervosa Register-based Severity Index evaluated 5 years after onset.

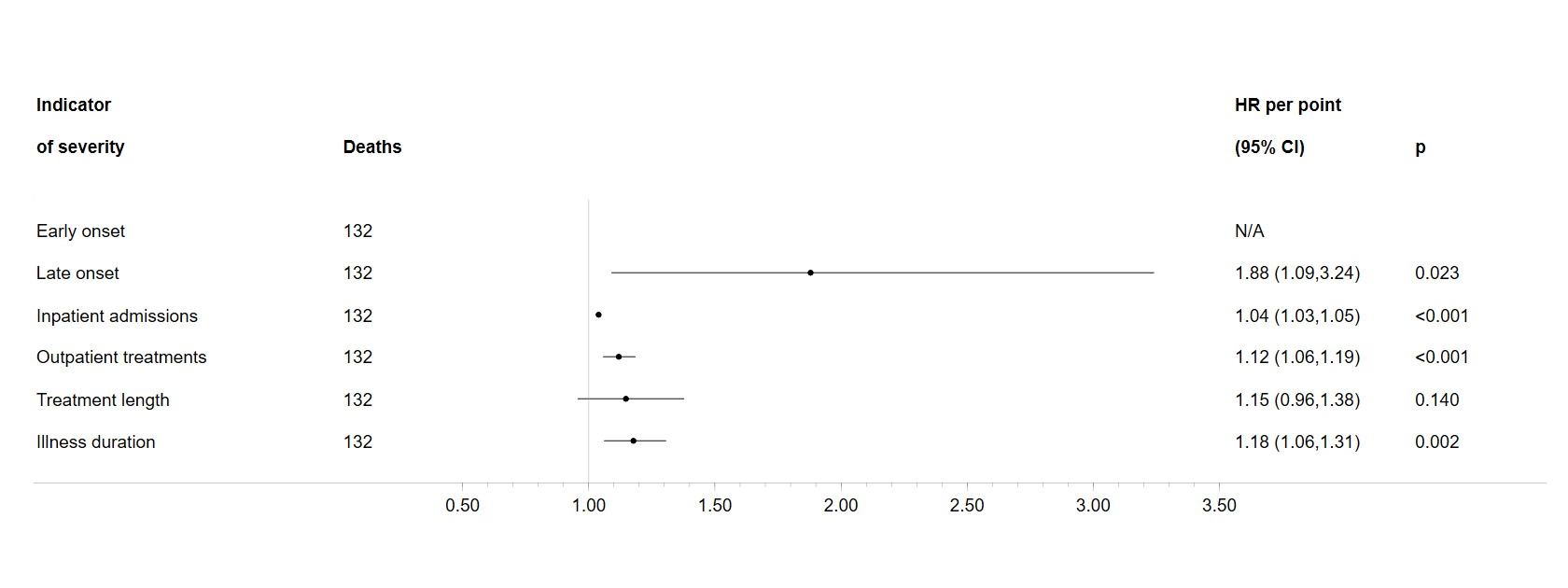

**Supplementary Figure 6.** Hazard ratios (HR) with 95% confidence intervals (CI) for overall mortality for severe anorexia nervosa cases compared to less-severe anorexia nervosa cases, defined as the top 20% vs. the bottom 80% of each of the individual severity variables (when possible) evaluated 5 years after onset.

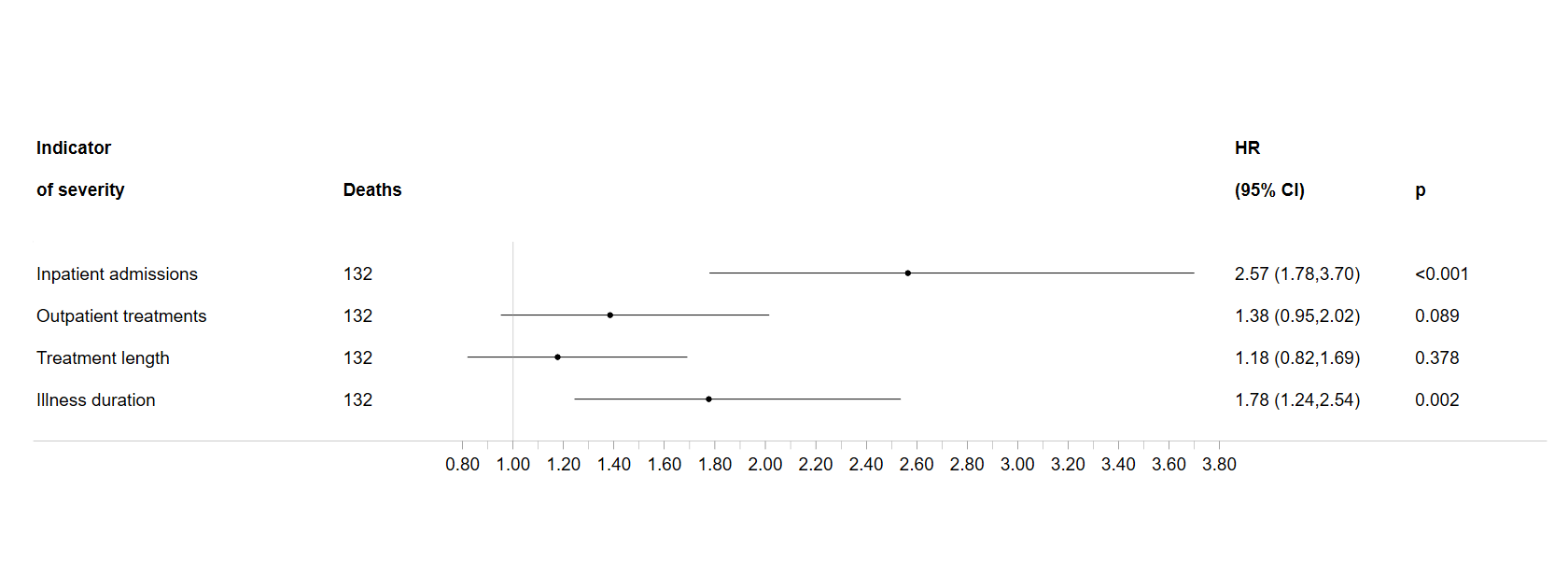

**Supplementary Figure 7.** Sex-stratified hazard ratios (HR) with 95% confidence intervals (CI) for overall mortality per one point increase of the Anorexia Nervosa Register-based Severity Index and for severe anorexia nervosa cases compared to less-severe anorexia nervosa cases evaluated 5 years after onset.

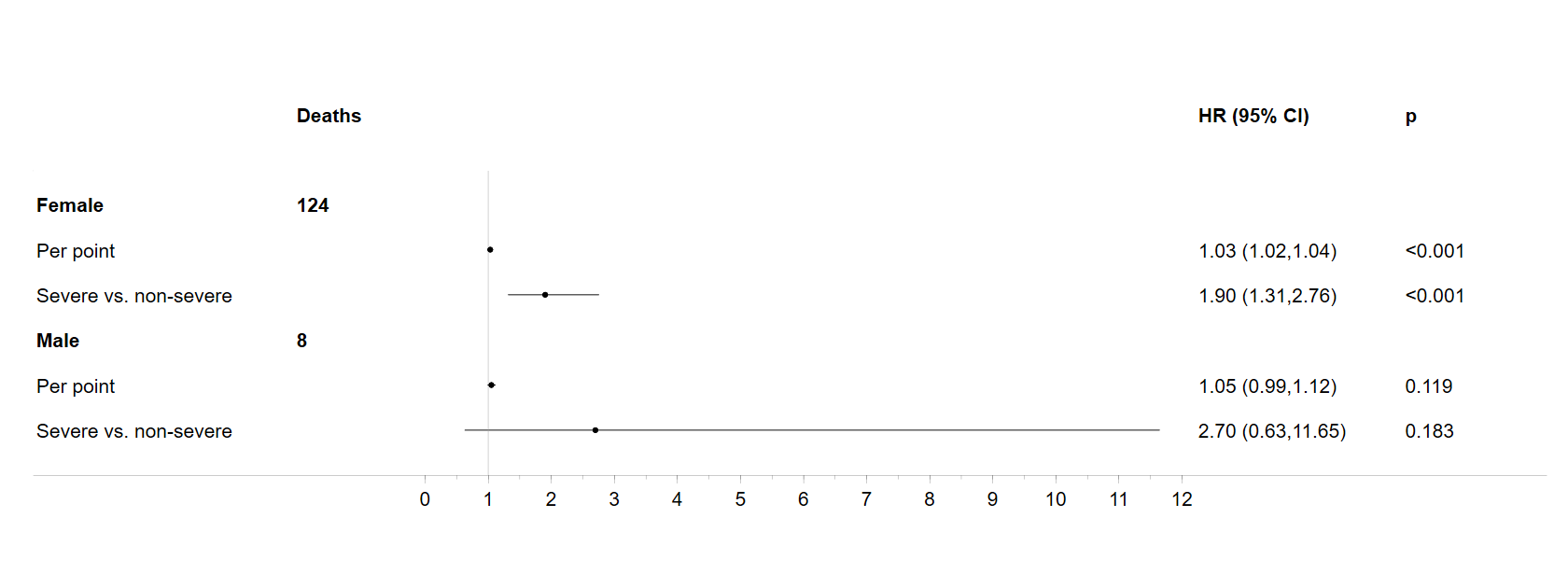

**Supplementary Figure 8.** Distribution of severity variables contributing to the combined Anorexia Nervosa Register-based Severity Index evaluated 5 years after AN onset using inpatient admission data only.

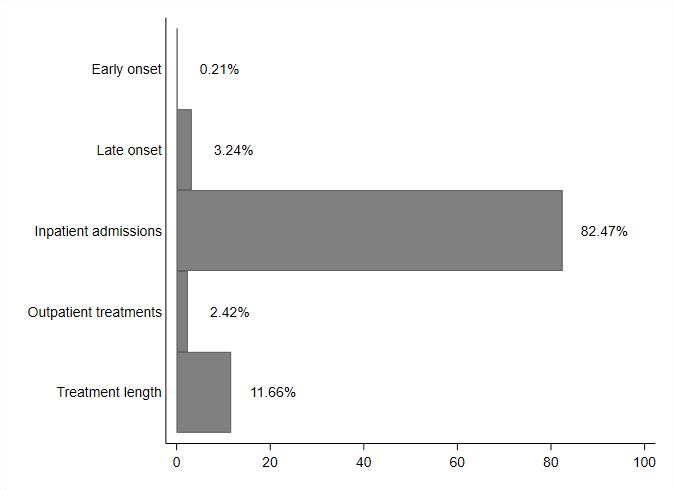

**Supplementary Figure 9.** Hazard ratios (HR) with 95% confidence intervals (CI) for overall and cause-specific mortality per one point increase of the Anorexia Nervosa Register-based Severity Index evaluated 5 years after onset using inpatient contacts only.

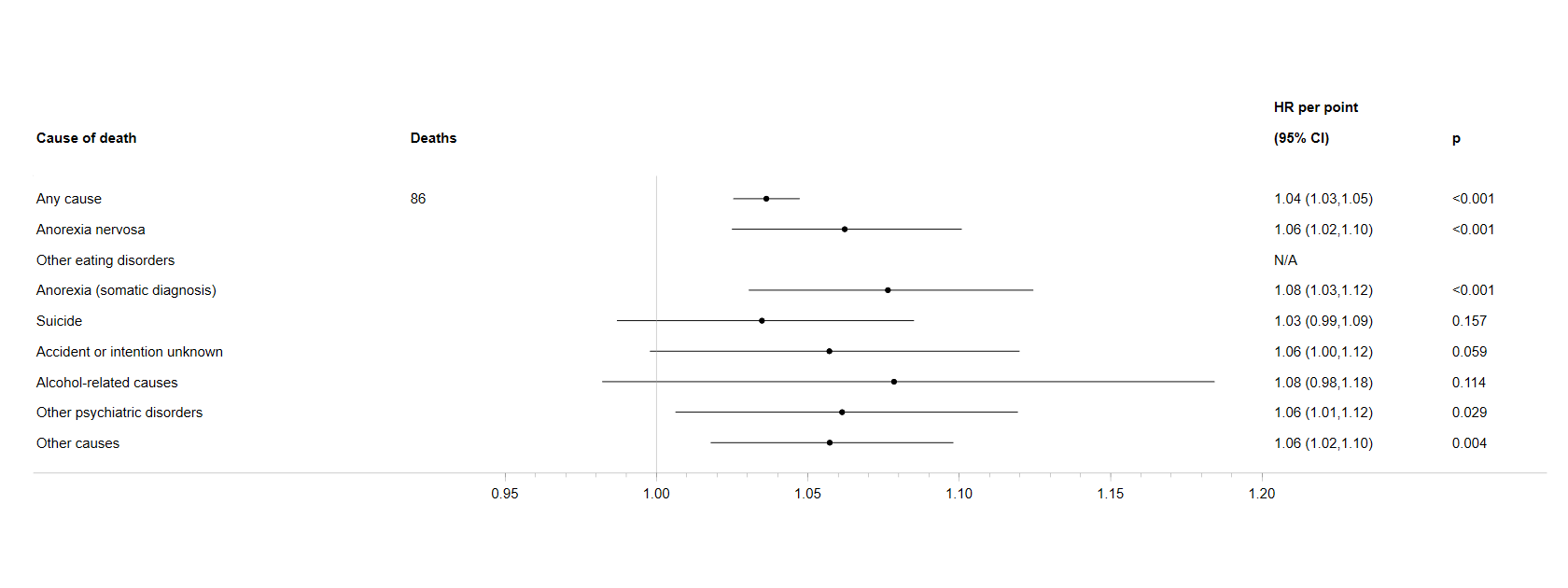

**Supplementary Figure 10.** Hazard ratios (HR) with 95% confidence intervals (CI) for overall and cause-specific mortality for severe anorexia nervosa cases compared to less-severe anorexia nervosa cases evaluated 5 years after onset using inpatient contacts only.

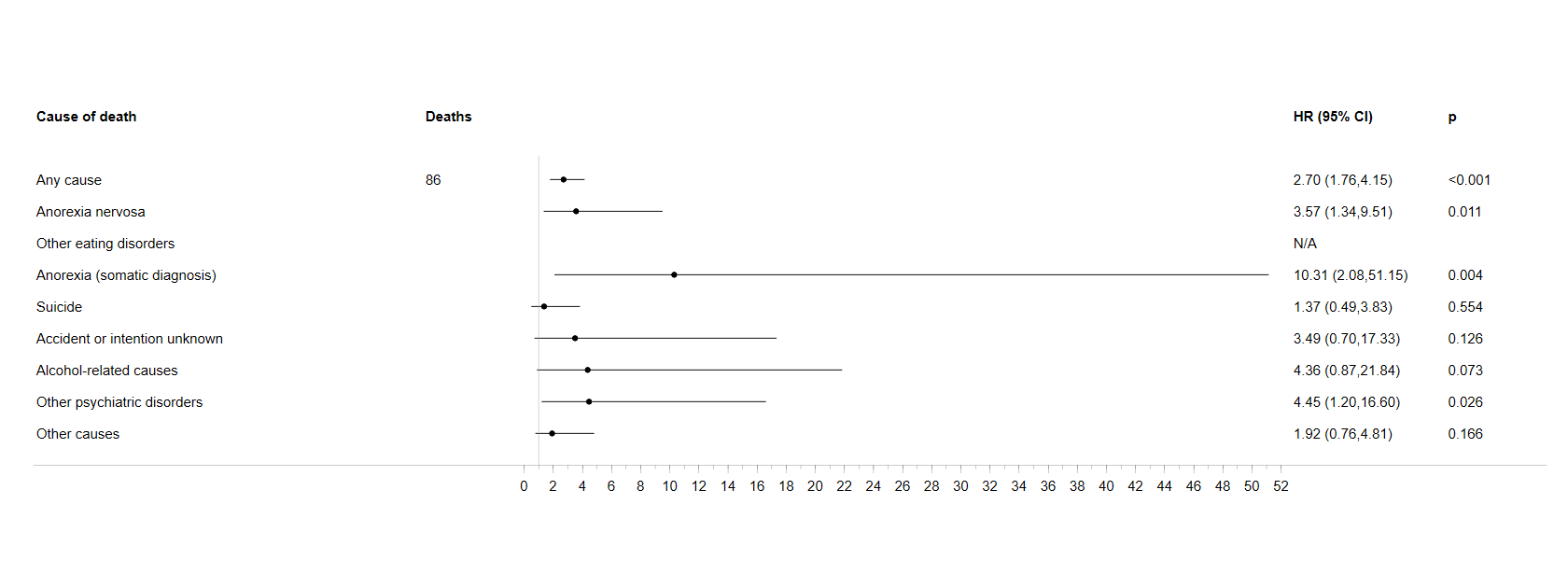
